# Accelerometry-Measured Physical Activity Predicts Dementia Onset in a National Cohort of Older Adults

**DOI:** 10.64898/2025.12.08.25341850

**Authors:** Matthew Ko, Ching-Hsuan Shirley Huang, Karl Brown, Kevin Vintch, Andrew Shutes-David, Katherine Wilson, Edmund Seto, Debby W. Tsuang

## Abstract

Identifying scalable, objective markers of dementia risk is essential for early prevention and intervention. Physical activity is a key modifiable risk factor, yet evidence from device-based measures in representative populations remains limited. Using nationally representative data from the National Health and Aging Trends Study (NHATS), we examined whether wrist-worn accelerometry predicts incident dementia in a cohort of 562 participants aged ≥ 65 years without dementia at baseline. Over one year of follow-up, higher physical activity levels were associated with lower risk of incident dementia (OR = 0.66, 95% CI [0.55 - 0.79]), even after adjusting for age, gender, and comorbidities (OR = 0.68, 95% CI [0.57 - 0.82]). Receiver operating characteristics (ROC) analyses indicate that accelerometry modestly outperforms self-reported physical capacity in predicting dementia onset. These findings support the use of wearable-derived activity metrics as a potential digital biomarker for dementia risk stratification in population and clinical settings.

## Background

Older adults exhibit a heightened vulnerability to dementia, functional disabilities, and other forms of cognitive impairment relative to younger populations [1]. With the prevalence of Alzheimer’s disease (AD) in the United States approaching 6 million and projected to reach 14 million by 2050, the associated societal burden—including impacts on individuals, caregivers, and healthcare infrastructure—is expected to escalate substantially.

Studies have found that physical activity is a major modifiable risk factor for cognitive impairment and dementia, suggesting that elderly individuals who lack consistent physical activity are at an increased risk of developing all forms of dementia [2, 3]. Furthermore, a higher proportion of elderly individuals with cognitive decline are more likely to be physically inactive compared to elderly individuals without cognitive impairment [4]. This suggests that there is a bi-directional relationship between cognitive function and physical activity, and that a lack of physical activity can worsen cognitive function and thus increase dementia risk [5].

Interventional studies such as FINGER and US-POINTER, which investigate the effects of physical activity on cognitive health, predominantly recruited healthy older adults [6, 7]. As a result, their findings may not be fully generalizable to the broader study population, particularly those with existing medical conditions, dementia or limited mobility. These recruitment biases limit the generalizability of the findings to a more vulnerable or diverse aging population.

Most observational studies examining the relationship between physical activity and dementia risk rely on self-reported measures of physical activity, which may introduce bias and limit the accuracy of the findings [8]. Thus, observational studies that utilize objectively measured physical activity such as accelerometry to investigate dementia risk improve the accuracy of the exposure assessment. Huang and colleagues utilized the UK Biobank to examine the association between physical activity metrics, such as daily step count, and the risk of incident dementia [9]. They found that a higher number of steps was associated with a decreased risk for all-cause dementia. Moreover, previous studies that have utilized actigraphy to study dementia are small feasibility studies [10], have focused on actigraphy-based sleep quality metrics [11], and combined physical activity and sleep metrics to assess dementia risk [12]. However, the association between quantifiable actigraphy metrics of physical activity and incident dementia risk has not been examined in a nationally representative sample of elderly individuals.

In the current study, we analyzed wrist-worn accelerometry data collected from a nationally representative sample of 562 older adults in US from the National Health and Aging Trends Study (NHATS), to quantify the association between physical activity and incident dementia. We aim to investigate physical activity as a key modifiable risk factor for reducing dementia risk in the elderly population [13]. By further analyzing these modifiable risk factors, and by leveraging the increasing ubiquity of wearable sensors, there is potential to improve cognitive health outcomes and quality of life for elderly individuals in the US [14, 15, 16].

## Methods

### Data Source and Study Population

The NHATS is a nationally representative longitudinal cohort initiated in 2011, enrolling Medicare beneficiaries aged 65 years and older in the US. It was established to track late-life functioning disability, and health among older adults. Annual in-person interviews and supplemental assessments provide longitudinal data on social, behavioral, and medical factors relevant to aging [17]. Round 11 data (collected in 2021) was the first wave to include adjudicated dementia status and collected actigraphy data (collected in 2022) for incident dementia outcomes.

### Dementia Classification

Dementia status in NHATS was adjudicated using established criteria [18]. Participants were classified as having *probable* dementia if they met at least one of the following criteria: (1) a self- or proxy-report of a physician diagnosis of dementia or AD; (2) an AD8 score ≥2 [19]; or (3) cognitive performance ≤1.5 standard deviations (SD) below the mean of at least two of three cognitive domains (memory, orientation, and executive function). *Possible* dementia was defined as scoring ≤1.5 SD below the mean in a single domain, while those negative across all domains were classified as having *no dementia*. Consistent with prior NHATS publications [20], participants with probable or possible dementia were combined into a single “dementia category” in the current analyses. Individuals classified as having dementia in Round 11 were excluded so that the current analyses would focus exclusively on incident dementia cases.

### Accelerometry Assessment

Accelerometry was first introduced in NHATS in 2019 following a pilot study demonstrating high participant compliance [21]. In Round 11, a random subsample was invited to wear an ActiGraph CentrePoint Insight Watch on the non-dominant wrist for eight consecutive days [22]. Of 944 eligible participants, 747 (86%) returned devices with usable data [21]. Devices recorded triaxial acceleration at 64 Hz in units of gravity (g). Participants were instructed to wear the watch continuously except during extended water-based activities. Detailed wear-time definitions, imputation procedures, and derived activity metrics have been described previously [21, 22]. Briefly, the accelerometry data was summarized into eight activity metrics: (1) total activity counts (ag11dtac), (2) minutes of active time (ag11dminact), (3) minutes of sedentary time (ag11minnon), (4) number of activity bouts (ag11dactnum), (5) average length of activity bouts (ag11dactlen), and maximum activity counts in (6) 10-minute (ag11dmax10), (7) 30-minute (ag11dmax30), and (8) 60-minute (ag11dmax60) intervals.

### Covariates

Covariates of the current analyses included demographic and health-related factors: age (70-74, 75-84, ≥85years), sex (male/female), and number of pre-existing health conditions. Comorbidities were based on self-reported physician diagnoses of heart disease, lung disease, myocardial infraction, hypertension, diabetes, cancer, stroke, arthritis, and osteoporosis [17]. Dementia was excluded from the comorbidity measure as it was the primary outcome.

### Statistical analysis

To reduce dimensionality, principal component analysis (PCA) was applied to the eight accelerometry metrics using the *svyprcomp* function in *R*, which accounts for the sample design in NHATS, including sample weights, strata, and clusters [21]. Metrics were centered and scaled using weighted means and standard deviations. The first principal component (PC1), which explained the greatest proportion of variance, was extracted and used as the primary predictor in the subsequent regression analyses of dementia outcomes.

The primary outcome was *incident dementia*, defined as new dementia classification (probable or possible) in Round 12 among participants without dementia in Round 11. Logistic regression models were fit using the svyglm function in R. We estimated both the crude model (eq. (1)) and models adjusted for covariates age, gender, and comorbidities (eq. (2)):

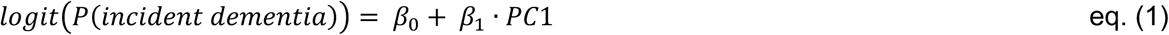

where the logit function *logit*(*P*(*incident dementia*)1 is the natural logarithm of the odds of incident dementia; *β*_0_ is the intercept, and *β*_1_ is the coefficient for PC1 of accelerometry variables, and

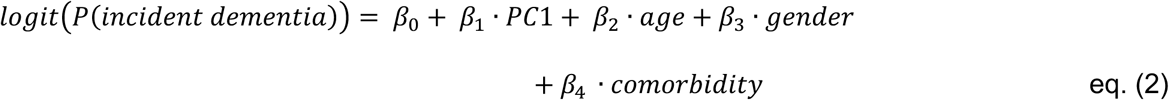

where the logit function *logit*(*P*(*incident dementia*)1 is the natural logarithm of the odds of the incident dementia; *β*_0_ is the intercept; *β*_1_ is the coefficient for PC1 of accelerometry variables; *β*_2_ is the coefficient of *age* variable (categorical, 0 = age 65 – 69; 1 = age 70 – 74; 2 = age 75 – 84; 3 = age 85+); *β*_3_ is the coefficient of the *gender* variable (0 = male; 1 = female), and *β*_4_ is the coefficient of the *comorbidity* variable (categorical, 0 = ≤ 4 comorbidities; 1 = >4 comorbidities). Model fit and significance of predictors were evaluated with Wald test, and odds ratios (OR) with 95% confidence intervals (CI) were reported for the association between accelerometry PC1 and incident dementia.

As a sensitivity analysis, the association between physical capacity (PC) scores and incident dementia was assessed. The PC score ranges from 0 to 4 and is based on the ability to perform nobility-related tasks. Briefly, the NHATS physical capacity (PC) score is based on mobility tasks involving walking and stair climbing [17]. For each task pair (walk 6 vs. 3 blocks; climb 20 vs. 10 stairs), participants able to perform the harder activity received 2 points and were not asked about the easier one. Those unable to perform the harder task but able to perform the easier one received 1 point. Scores across both pairs were summed, yielding a maximum PC score of 4. An adjusted sensitivity model including age, gender, and comorbidities was also assessed. Model performance was determined using the Area Under the Curve (AUC) of the Receiver Operating Characteristic (ROC) curve. All statistical analyses were performed using *R* version 4.2.3. The survey design was specified using the *svydesign* function of the *survey*package.

## Results

The selected cohort that was chosen was more closely skewed female (54.3%) (**Table 1**). However, of the individuals who developed incident dementia within the 1-year time frame, more were male (60.5%). Most participants identified as White, non-Hispanic (84.2%), with a smaller proportion identifying as Non-White (13.7%) and 2.1% not reporting race/ethnicity. Compared to previous literature of demographic characteristics of Alzheimer’s disease (AD), more women are typically affected than males [23, 24]. The overall incidence of dementia between Round 11 to 12 was 11% for men and 10% for women.

**Table 1.** Demographic characteristics of eligible participants who did or did not develop dementia from Round 11 to 12.

|  | No dementia<br>(N=519) | Incident dementia<br>(N=43) | Overall<br>(N=562) |
| --- | --- | --- | --- |
| **Age** |  |  |  |
| 70-74 | 162 (31.2%) | 9 (20.9%) | 171 (30.4%) |
| 75-84 | 283 (54.5%) | 22 (51.2%) | 305 (54.3%) |
| 85+ | 74 (14.3%) | 12 (27.9%) | 86 (15.3%) |
| **Race/Ethnicity <sup>1</sup>** |  |  |  |
| White, non-Hispanic | 442 (85.2%) | 31 (72.1%) | 473 (84.2%) |
| Non-White | 67 (12.9%) | 10 (23.3%) | 77 (13.7%) |
| Non-selected | 10 (1.9%) | 2 (4.6%) | 12 (2.1%) |
| **Gender** |  |  |  |
| Male | 231 (44.5%) | 26 (60.5%) | 257 (45.7%) |
| Female | 288 (55.5%) | 17 (39.5%) | 305 (54.3%) |
| **Comorbid conditions** |  |  |  |
| ≤4 | 474 (91.3%) | 39 (90.7%) | 513 (91.3%) |
| >4 | 45 (8.7%) | 4 (9.3%) | 49 (8.7%) |

<sup>1</sup> Non-White group includes Black, Hispanic, Asian, Native American, Pacific Islander, or multiple races; Non-selected indicates missing or refused responses.

Table 2 summarizes the cognitive assessments used by NHATS to evaluate cognitive and dementia status. Individuals who developed incident dementia scored significantly lower than those without dementia across all domains, including orientation (date recall and naming of the President and Vice President), executive function (clock-drawing score), and memory (immediate and delayed word recall; p< 0.001 for all). The AD8 measure was not compared statistically due to near-complete missingness (>97%) in both groups, as it is administered only to proxy respondents that would possibly cause this [17].

**Table 2.** Cognitive assessments result in Round 12 by dementia status.

|  | No dementia<br>(N=519) | Incident dementia<br>(N=43) | Overall<br>(N=562) | p-value <sup>1</sup> |
| --- | --- | --- | --- | --- |
| AD8 |  |  |  |  |
| Score ≥ 2 | 0 (0%) | 1 (2.3%) | 1 (0.2%) | N/A <sup>2</sup> |
| Score <2 | 1 (0.2%) | 0 (0%) | 1 (0.2%) |  |
| Missing | 518 (99.8%) | 42 (97.7%) | 560 (99.6%) |  |
| Date Items Recalled |  |  |  |  |
| Mean (SD) | 3.78 (0.48) | 2.98 (1.22) | 3.72 (0.60) | <0.001 |
| Median | 4.00 | 3.50 | 4.00 |  |
| [Min, Max] | [1.00, 4.00] | [0.00, 4.00] | [0.00, 4.00] |  |
| Missing | 0.00 (0.00%) | 1.00 (2.33%) | 1.00 (0.2%) |  |
| President/Vice President Names |  |  |  |  |
| Mean (SD) | 3.50 (0.92) | 1.90 (1.49) | 3.38 (1.06) | <0.001 |
| Median | 4.00 | 2.00 | 4.00 |  |
| [Min, Max] | [0.00, 4.00] | [0.00, 4.00] | [0.00, 4.00] |  |
| Missing | 0.00 (0.00%) | 1.00 (2.33%) | 1 (0.2%) |  |
| Orientation: Sum of Date Recall and President/Vice President/Vice Names |  |  |  |  |
| Mean (SD) | 7.28 (1.07) | 4.88 (2.20) | 7.10 (1.34) | <0.001 |
| Median | 8.00 | 5.00 | 8.00 |  |
| [Min, Max] | [4.00, 8.00] | [1.00, 8.00] | [1.00, 8.00] |  |
| Missing | 0.00 (0.00%) | 1.00 (2.33%) | 1 (0.2%) |  |
| **Executive: Clock Drawing Score** |  |  |  |  |
| Mean (SD) | 4.25 (0.88) | 3.02 (1.49) | 4.16 (0.99) | <0.001 |
| Median | 4.00 | 3.00 | 4.00 |  |
| [Min, Max] | [2.00, 5.00] | [0.00, 5.00] | [0.00, 5.00] |  |
| **Immediate Word Recall** |  |  |  |  |
| Mean (SD) | 5.47 (1.45) | 2.64 (1.56) | 5.26 (1.64) | <0.001 |
| Median | 5.00 | 2.50 | 5.00 |  |
| [Min, Max] | [2.00, 10.00] | [0.00, 7.00] | [0.00, 10.00] |  |
| Missing | 0.00 (0.00%) | 1.00 (2.33%) | 1.00 (0.2%) |  |
| **Delayed Word Recall** |  |  |  |  |
| Mean (SD) | 4.25 (1.82) | 1.10 (1.56) | 4.01 (1.98) | <0.001 |
| Median | 4.00 | 0.00 | 4.00 |  |
| [Min, Max] | [0.00, 9.00] | [0.00, 6.00] | [0.00, 9.00] |  |
| Missing | 0.00 (0.00%) | 1.00 (2.33%) | 1.00 (0.2%) |  |
| **Memory: Immediate and Delayed Word Recall** |  |  |  |  |
| Mean (SD) | 9.72 (2.97) | 3.74 (2.85) | 9.27 (3.35) | <0.001 |
| Median | 10.00 | 3.00 | 9.00 |  |
| [Min, Max] | [4.00, 18.00] | [0.00, 12.0] | [0.00, 18.0] |  |
| Missing | 0.00 (0.00%) | 1.00 (2.33%) | 1.00 (0.2%) |  |

<sup>1</sup> p-values were obtained from design-based Wilcoxon rank-sum tests using the *svyranktest* function in the R survey package, accounting for NHATS sampling weights, strata, and clusters.
<sup>2</sup> Not tested due to near-complete missingness (>97.0%) in both groups.

Table 3 summarizes the accelerometry-derived physical activity metrics from NHATS. Across all measures, individuals who developed incident dementia were significantly less active than those without dementia (p< 0.01 for all). The incident dementia group demonstrated lower activity counts, fewer active minutes per day, and fewer activity bouts, as well as shorter average bout durations. They also exhibited lower maximum activity counts over 10-, 30-, and 60-minute intervals, indicating reduced intensity and duration of movement throughout the monitoring period.

**Table 3.** Accelerometry metrics by dementia status.

|  | **No dementia**<br>**(N=519)** | **Incident**<br>**dementia**<br>**(N=43)** | **Overall**<br>**(N=562)** | **p-value<sup>1</sup>** |
| --- | --- | --- | --- | --- |
| **Total Activity Counts (ag11dtac)** |  |  |  |  |
| Mean ± SD | $1.8 \times 10^6 \pm 6.1 \times 10^5$ | $1.4 \times 10^6 \pm 5.4 \times 10^5$ | $1.8 \times 10^6 \pm 6.2 \times 10^5$ | <0.001 |
| Median | $1.8 \times 10^6$ | $1.4 \times 10^6$ | $1.7 \times 10^6$ | |
| [Min, Max] | $[2.0 \times 10^4, 4.3 \times 10^6]$ | $[3.1 \times 10^5, 2.9 \times 10^6]$ | $[2.0 \times 10^4, 4.3 \times 10^6]$ | |
| **Minutes Active per Day (ag11dminact)** |  |  |  |  |
| Mean ± SD | $3.6 \times 10^2 \pm 1.2 \times 10^2$ | $2.9 \times 10^2 \pm 1.2 \times 10^2$ | $3.6 \times 10^2 \pm 1.2 \times 10^2$ | <0.001 |
| Median | $3.6 \times 10^2$ | $2.8 \times 10^2$ | $3.6 \times 10^2$ | |
| [Min, Max] | $[0.00, 7.4 \times 10^2]$ | $[2.1 \times 10^1, 6.5 \times 10^2]$ | $[0.00, 7.4 \times 10^2]$ | |
| **Minutes Non-Active per Day (ag11dminnon)** |  |  |  |  |
| Mean ± SD | $1.1 \times 10^3 \pm 1.2 \times 10^2$ | $1.2 \times 10^3 \pm 1.2 \times 10^2$ | $1.1 \times 10^3 \pm 1.2 \times 10^2$ | <0.001 |
| Median | $1.1 \times 10^3$ | $1.2 \times 10^3$ | $1.1 \times 10^3$ | |
| [Min, Max] | $[7.0 \times 10^2, 1.4 \times 10^3]$ | $[7.9 \times 10^2, 1.4 \times 10^3]$ | $[7.0 \times 10^2, 1.4 \times 10^3]$ | |
| **Active Bouts per Day<sup>2</sup>(ag11dactnum)** |  |  |  |  |
| Mean ± SD | $9.0 \times 10^1 \pm 1.9 \times 10^1$ | $8.3 \times 10^1 \pm 2.5 \times 10^1$ | $9.0 \times 10^1 \pm 2.0 \times 10^1$ | <0.01 |
| Median | $9.1 \times 10^1$ | $8.5 \times 10^1$ | $9.0 \times 10^1$ | |
| [Min, Max] | $[0.0, 1.4 \times 10^2]$ | $[1.6 \times 10^1, 1.5 \times 10^2]$ | $[0.0, 1.5 \times 10^2]$ | |
| **Mean Length Active Bouts (ag11dactlen)** |  |  |  |  |
| Mean ± SD | $4.0 \pm 1.2$ | $3.5 \pm 1.0$ | $4.0 \pm 1.2$ | <0.01 |
| Median | 3.9 | 3.5 | 3.9 |  |
| [Min, Max] | $[0.0, 1.0 \times 10^1]$ | $[1.4, 5.4]$ | $[0.0, 1.0 \times 10^1]$ | |

|  | No dementia<br>(N=519) | Incident<br>dementia<br>(N=43) | Overall<br>(N=562) | p-value<sup>1</sup> |
| --- | --- | --- | --- | --- |
| **Max Activity Counts in 10 Consecutive Min (ag11dmax10)** |  |  |  |  |
| Mean ± SD | 6.5 x 10<sup>3</sup> ± 1.7 x 10<sup>3</sup> | 5.5 x 10<sup>3</sup> ± 1.7 x 10<sup>3</sup> | 6.4 x 10<sup>3</sup> ± 1.7 x 10<sup>3</sup> | <0.001 |
| Median | 6.4 x 10<sup>3</sup> | 5.6 x 10<sup>3</sup> | 6.4 x 10<sup>3</sup> |  |
| [Min, Max] | [4.1 x 10<sup>2</sup>, 2.0 x 10<sup>4</sup>] | [1.6 x 10<sup>3</sup>, 1.1 x 10<sup>4</sup>] | [4.1 x 10<sup>2</sup>, 2.0 x 10<sup>4</sup>] |  |
| **Max Activity Counts in 30 Consecutive Min (ag11dmax30)** |  |  |  |  |
| Mean ± SD | 5.0 x 10<sup>3</sup> ± 1.4 x 10<sup>3</sup> | 4.1 x 10<sup>3</sup> ± 1.4 x 10<sup>3</sup> | 5.0 x 10<sup>3</sup> ± 1.5 x 10<sup>3</sup> | <0.001 |
| Median | 4.9 x 10<sup>3</sup> | 4.3 x 10<sup>3</sup> | 4.9 x 10<sup>3</sup> |  |
| [Min, Max] | [1.5 x 10<sup>2</sup>, 1.6 x 10<sup>4</sup>] | [1.1 x 10<sup>3</sup>, 9.0 x 10<sup>3</sup>] | [1.5 x 10<sup>2</sup>, 1.6 x 10<sup>4</sup>] |  |
| **Max Activity Counts in 60 Consecutive Min (ag11dmax60)** |  |  |  |  |
| Mean ± SD | 4.1 x 10<sup>3</sup> ± 1.3 x 10<sup>3</sup> | 3.4 x 10<sup>3</sup> ± 1.3 x 10<sup>3</sup> | 4.0 x 10<sup>3</sup> ± 1.3 x 10<sup>3</sup> | <0.001 |
| Median | 4.0 x 10<sup>3</sup> | 3.3 x 10<sup>3</sup> | 4.0 x 10<sup>3</sup> |  |
| [Min, Max] | [8.4 x 10<sup>3</sup>, 1.2 x 10<sup>4</sup>] | [8.9 x 10<sup>2</sup>, 8.3 x 10<sup>3</sup>] | [8.4 x 10<sup>3</sup>, 1.2 x 10<sup>4</sup>] |  |

<sup>1</sup> p-values were obtained from design-based Wilcoxon rank-sum tests using the *svyranktest* function in the R survey package, accounting for NHATS sampling weights, strata, and clusters.
<sup>2</sup> Active Bouts defined as an uninterrupted sequence of ≥1 active minutes (minutes with >1853 activity counts).<sup>21</sup> [21].

**Figure 1** presents the correlation matrix for accelerometry metrics across dementia and non-dementia groups, including the correlations of each variable with PC1, which serves as a summary measure of physical activity. In the dementia group, variables associated with higher physical activity levels, such as daily total activity counts (ag11dtac), number of active bouts per day (ag11dactnum), and maximum activity counts over 10, 30, and 30 consecutive minutes (ag11dmax10, ag11dmax30, ag11dmax60), show stronger internal correlations than in the non-dementia group. However, the distribution of these metrics reflects the expected trend, with lower overall activity levels in the dementia group. Metrics indicating less activity, such as the number of non-active minutes per day (ag11dminnon), have stronger correlations within the dementia group, showing the differences in activity patterns between groups.

**Figure 1.**
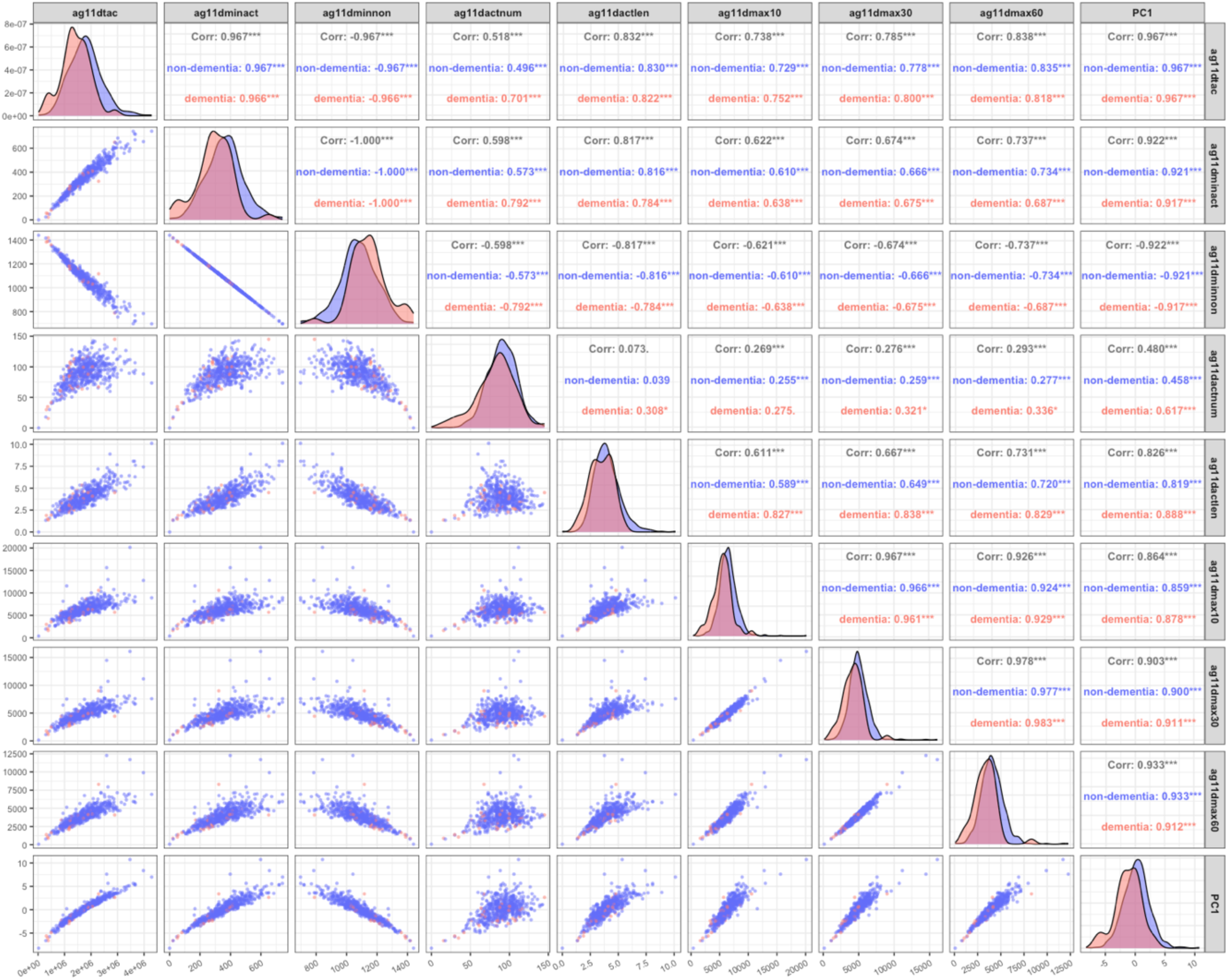
Correlation matrix of the accelerometry variables and PC1 for the dementia and non-dementia group. Variable description: ag11dtac: daily total activity counts; ag11dminact: number of minutes active per day; ag11dminnon: number of minutes non-active per day; ag11dactnum: number of active bouts per day; ag11dactlen: mean length of active bouts; ag11dmax10: max activity counts in 10 consecutive minutes; ag11dmax30: max activity counts in 30 consecutive minutes; ag11dmax60: max activity counts in 60 consecutive minutes

PC1, representing a composite of physical activity levels, is positively correlated with all accelerometry variables related to higher physical activity level, suggesting PC1 captures the general intensity of physical activity level. **Figure S1** shows that PC1 explains approximately 70% of the variance, whereas **Table S1** provides the loading of each variable on PC1, PC2, and PC3, with PC1 showing positive loadings across all metrics associated with higher physical activity level and negative loadings for metric related to lower physical activity level (ag11dminnon).

**Table 4** shows the associations between the PCA-derived physical activity (PC1), demographic factors, comorbidity burden, and incident dementia. Higher physical activity levels, as indicated by PC1, were significantly associated with lower dementia risk in both the crude model (OR = 0.66, 95% CI: 0.55 – 0.79, p < 0.001) and the adjusted model (OR = 0.68, 95% CI: 0.57 – 0.82, p < 0.001), independent of age, gender, and comorbidities. For age, there was a non-significant trend of increased dementia risk with older age groups, with adjusted odds ratios of 1.34 (95% CI: 0.58 – 3.09) for ages 75 – 84 and 2.19 (95% CI: 0.89 – 5.42) for ages 85+. Female gender was associated with a lower but non-significant risk (OR = 0.58, 95% CI: 0.32 – 1.07, p = 0.09), while comorbidity burden showed no significant effect on dementia risk (OR = 1.08, 95% CI: 0.23 – 4.98, p = 0.91).

**Table 4.** Association between accelerometry and incident dementia.

| Variables | Crude model <sup>1</sup> |  | Adjusted model <sup>1</sup> |  |
| --- | --- | --- | --- | --- |
|  | OR (95% CI) | *p* | OR (95% CI) | *p* |
| *PC1 (accelerometry)* <sup>2</sup> | 0.66 (0.55 – 0.79) | <0.001 | 0.68 (0.57 – 0.82) | <0.001 |
| *Age (years)* |  |  |  |  |
| 70 – 74 | - | - | 1.00 |  |
| 75 – 84 | - | - | 1.34 (0.58 – 3.09) | 0.45 |
| 85+ | - | - | 2.19 (0.89 – 5.42) |  |
| *Gender* |  |  |  |  |
| Male | - | - | 1.00 |  |
| Female | - | - | 0.58 (0.32 – 1.07) | 0.09 |
| *Comorbidity conditions* <sup>3</sup> |  |  |  |  |
| ≤4 | - | - | 1.00 |  |
| >4 | - | - | 1.08 (0.23 – 4.98) | 0.91 |

Definition of abbreviations: OR: odds ratio; CI: confidence interval.
<sup>1</sup> Accounted for survey design, including sample weights, strata, and clusters.
<sup>2</sup> Combined accelerometry score based on principal component analysis component 1 (PC1). A higher score indicates a higher physical activity level.
<sup>3</sup> Numbers of existing and/or pre-existing comorbidities.

**Table S2** and **Figure S2** show the sensitivity analysis results using physical capacity (PC) as the predictor of dementia risk. Similar to the main modeling results, higher PC scores were associated with a lower risk of dementia in both the crude (OR = 0.69, 95% CI: 0.54 – 0.89, p < 0.001) and adjusted models (OR = 0.64, 95% CI: 0.48 – 0.85, p <0.01). Female gender was also associated with lower dementia risk (OR = 0.32, 95% CI: 0.16 – 0.64, p<0.001), while age and comorbidities showed no significant associations.

The ROC cures in **Figure S2** indicate similar predictive accuracy between models, with AUCs of 0.697 for PC1 (accelerometry) and 0.691 for PC (physical capacity) in the adjusted models, supporting the robustness of the findings.

## Discussion

In this nationally representative cohort of older adults, higher objectively measured physical activity was associated with a reduced risk of developing dementia within one year. This relationship persisted even after adjustment for age, sex and various comorbidities. The results suggest that improving physical activity levels can be an effective way of reducing dementia risk in a vulnerable yet growing population.

It is well documented that incident dementia risk is correlated with various modifiable risk factors. Alongside physical inactivity, factors such as social isolation, depression, smoking, and diabetes have been identified as important contributors to increased dementia risk [25]. Prior studies have demonstrated that older adults who maintain good physical health, limit or abstain from alcohol and smoking, and consume a healthy, balanced diet are at a lower risk of developing dementia and related cognitive conditions [25, 26, 27]. Current evidence from both large observational studies and meta-analyses of these studies suggests that adults who are physically active are less likely to develop dementia when compared to individuals who are less active.

The Finnish Geriatric Intervention Study to Prevent Cognitive Impairment and Disability (FINGER) provided landmark randomized controlled evidence supporting the role of lifestyle modification in mitigating cognitive decline and subsequent disease progression [6]. The trial recruited 1260 participants aged 60-77 years who were at risk for dementia and implemented a two-year multi-domain intervention targeting physical activity, diet, cognitive training, and vascular and metabolic risk management. Compared to a control group receiving standardized health advice, the interventional arm participants demonstrated significant improvements in cognitive performance. Importantly, these benefits were consistent across subgroups defined by sex, education and income [6]. Findings from FINGER promote the concept of physical activity, as a part of a broader intervention towards a healthier lifestyle, can yield cognitive benefit in older individuals who may exhibit other dementia risk factors.

The U.S. Study to Protect Brain Health Through Lifestyle Intervention to Reduce Risk (US POINTER) is a randomized controlled trial modified after the FINGER trial. The large, multisite trial aimed to test whether structured lifestyle interventions can preserve cognitive function in older adults at risk for developing dementia [7]. These multidomain interventions focused on further understanding of four identified areas of concern, including physical activity, nutrition, cognitive/social engagement, and vascular management. Importantly, the US POINTER demonstrated that structured, higher intensity exercise can improve cognition. The US POINTER trial provides key translation evidence that lifestyle adjustment, including reducing vascular risk and improving physical activity, can maintain cognition in older adults. Complementing these findings, the PREVENT dementia program provided a multi-center prospective data set that found mid-life intervention with the early signs of neurodegenerative disease identification [28]. Focused on individuals aged 40-59 years, the PREVENT study examined modifiable and non-modifiable determinants of dementia risk. Findings have shown that midlife modifiable risk factors are heavily associated with markers of small vessel disease in cognitively healthy adults, which is known to play a key role in developing dementia at a later age [29]. These findings indicate that addressing modifiable risk factors earlier in the course of life, such as physical activity, is helpful in reducing dementia risk later in life.

While the evidence of physical activity and risk of developing dementia is well noted, there is a notable gap in understanding the underpinnings of physical activity and dementia risk. Previous cohort studies, such as the Amsterdam and Framingham studies, have compared various risk factors but did not address the effect of physical activity. The Amsterdam Study published by van der Flier et al. focused on primarily the diagnosis and clinical manifestation of dementia patients [30]. There was limited discussion on the effects of physical activity in reducing incident dementia risk. Similarly, Satizabal et al. investigated the incidence of dementia in a cohort of individuals followed by the Framingham Heart study [31]. Results of the study showed that the incidence of dementia has decreased from a 30-year period. However, there were no reported outcomes on accelerometry metrics. Similarly to the present study, Son et al. compared accelerometry-measured patterns of sedentary behaviors and functional status between individuals using the NHATS database [32]. They found that more time spent in sedentary behavior was associated with higher cognitive function, stronger hand grip strength and short physical battery performance, and less difficulties with activities of daily living [32].

The overall incidence of dementia increases with age: incidence of 5.3% in those 70-74 years old, 7.0% in those 75-84 years old, and 13.0% in those over 85 years old. These rates are comparable to estimates previously reported: 5.7-6.5% in those 75-84 years old, and 13.1% in those over 85 years old [33]. Surprisingly, we found that men had a slightly higher incidence rate than women, 11.0% versus 10.0%, respectively. However, these differences fluctuated across the rounds, with four rounds men observed as having higher incidence, 3 rounds with women observed having higher incidence and 3 other rounds with equal incidence rates between the sexes. According to the 2015 Medicare data reported to the CDC, the incidence of dementia was similar in both sexes (women 3.4% and men 3.5%). However, the 2021 Medicare data showed that while overall incidence of dementia decreased more dramatically in women (2.6%) versus in men (2.9%) [34]. The NHATS data likely reflects these changes.

This study provides an association between objectively measured physical activity levels and dementia, and our findings provide a foundation for future studies to investigate other actigraphy metrics and physical function measurements. A key strength of this study lies in the use of a nationally representative sample in the U.S., allowing for generalization of findings to the broader U.S. elderly adult population. Similar to the other rounds, the annual incidence of dementia is approximately 10.0%. Unexpectedly, in this round, the incidence of dementia is higher in males versus females. Additionally, NHATS includes oversampling of very old age groups (e.g. 90+) and black non-Hispanic individuals, thus improving the representation of subgroups often underrepresented in aging research. The use of accelerometry data provides an objective measure of physical activity, reducing self-report bias, and improving measurement precision.

This study had several limitations. Namely, the number of individuals who developed incident dementia (n=43) was limited, and we were only able to examine the incidence of dementia over 1 year. These limitations can be addressed in future years when NHATS releases new rounds of actigraphy and cognitive assessment data. Additionally, the possibility of reverse causation cannot be ruled out; individuals with early or preclinical dementia may voluntarily reduce their physical activity prior to their clinical diagnosis. Additional studies are necessary to investigate the direction of the association between physical activity and dementia.

## Data Availability

All data produced in the present study are available upon reasonable request to the authors.

https://www.nhats.org

## Author Contributions

Conceptualization: ES. Methodology: SH, KB, KW, ES, DT. Formal analysis: MK, SH, ES. Writing: MK, SH, KB, AS, KW, ES, DT. Supervision and Project administration: ES, DT.

## Acknowledgments

We thank Dr. Jennifer Schrack for assistance with accelerometry data in the NHATS.

## Datasets/Data Availability Statement

All data used in these analyses were obtained from public data files available on the NHATS website:

