## Supplemental Info for "Accelerometry-Measured Physical Activity Predicts Dementia Onset in a National Cohort of Older Adults"

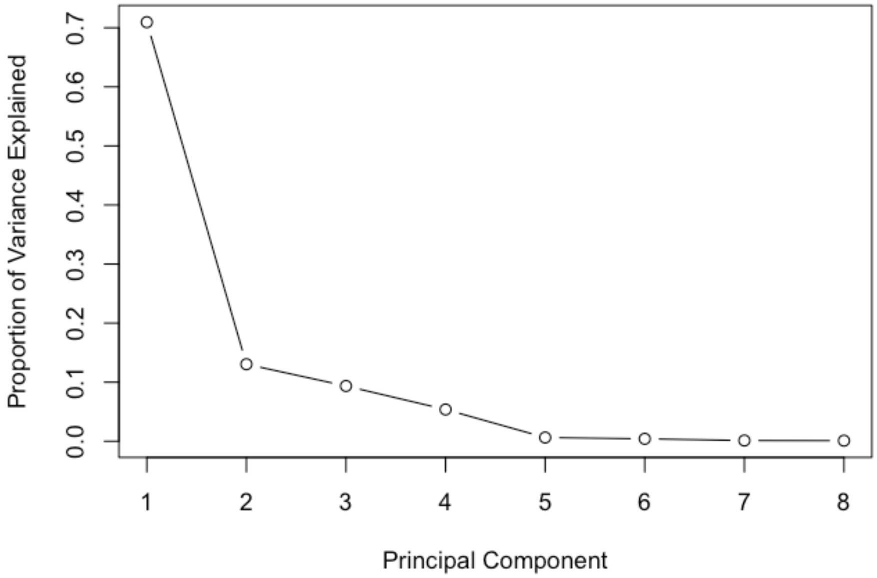


**Figure S1. Screen plot for principal component analysis on accelerometry metrics.**

**Table S1. Loading values of PCA for the three main components (PC1 – PC3) for accelerometry.**

| Variables | PC1 | PC2 | PC3 |
|---|---:|---:|---:|
| ag11dtac | 0.407 | 0.124 | -0.043 |
| ag11dminact | 0.392 | 0.269 | -0.060 |
| ag11dminnon | -0.250 | -0.454 | 0.632 |
| ag11dactnum | 0.224 | 0.629 | 0.632 |
| ag11dactlen | 0.356 | -0.175 | -0.336 |
| ag11dmax10 | 0.371 | -0.339 | 0.223 |
| ag11dmax30 | 0.384 | -0.314 | 0.163 |
| ag11dmax60 | 0.395 | -0.257 | 0.082 |

Variable description: ag11dtac: daily total activity counts; ag11dminact: number of minutes active per day; ag11dminnon: number of minutes non-active per day; ag11dactnum: number of active bouts per day; ag11dactlen: mean length of active bouts; ag11dmax10: max activity counts in 10 consecutive minutes; ag11dmax30: max activity counts in 30 consecutive minutes; ag11dmax60: max activity counts in 60 consecutive minutes.

**Table S2. Sensitivity analysis: summary of the modeling results using physical capacity score (PC) as the main predictor.**

| Variables | | Crude model [1] | | Adjusted model [1] | |
|---|---|---|---|---|---|
| | | OR (95% CI) | *p* | OR (95% CI) | *p* |
| *PC score* | | 0.69 (0.54 – 0.89) | <0.001 | 0.64 (0.48 – 0.85) | <0.01 |
| *Age (years)* | | | | | |
| | 70 – 74 | - | - | 1.00 | 0.52 |
| | 75 – 84 | - | - | 1.35 (0.59 – 3.07) | |
| | 85+ | - | - | 2.18 (0.80 – 5.92) | |
| *Gender* | | | | | |
| | Male | - | - | 1.00 | <0.001 |
| | Female | - | - | 0.32 (0.16 – 0.64) | |
| *Comorbidity conditions* [2] | | | | | |
| | ≤4 | - | - | 1.00 | 0.61 |
| | >4 | - | - | 0.74 (0.22 – 2.50) | |

Definition of abbreviations: OR: odds ratio; CI: confidence interval.

[1] Accounted for survey design, including sample weights, strata, and clusters.

[2] Numbers of existing and/or pre-existing comorbidities.

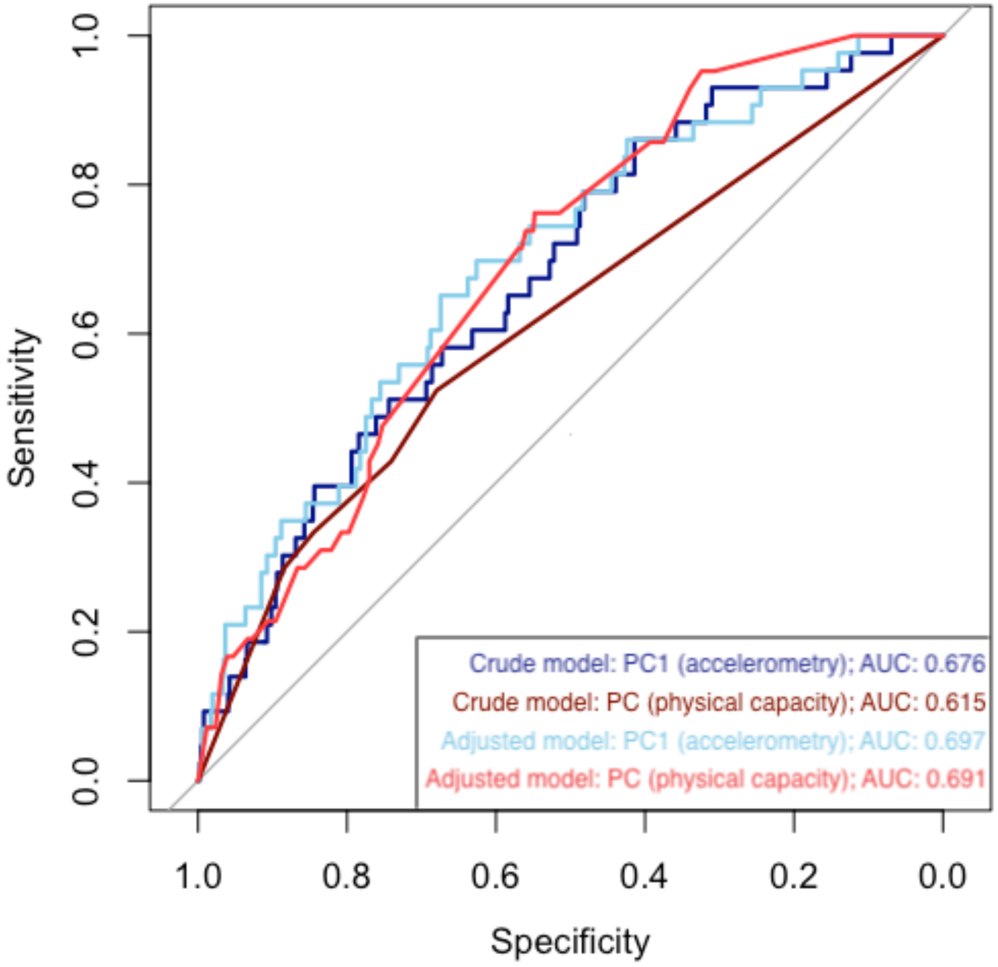


**Figure S2. ROC curves and AUC of the evaluated models.**
